# Parkinson’s disease-related motor and non-motor symptoms are not more prevalent in the Lancaster Amish

**DOI:** 10.1101/2020.04.13.20064337

**Authors:** Michael D.F. Goldenberg, Xuemei Huang, Honglei Chen, Lan Kong, Teodor T. Postolache, John W. Stiller, Katherine A. Ryan, Mary Pavlovich, Toni I. Pollin, Alan R. Shuldiner, Richard B. Mailman, Braxton D. Mitchell

**Author notes:** Corresponding Authors: Xuemei Huang, M.D., Ph.D., Department of Neurology, Penn State Hershey Medical Center, 500 University Dr., H-037, Hershey, PA 17033-0850, Braxton D. Mitchell, Ph.D., Program for Personalized and Genomic Medicine, Division of Endocrinology, Diabetes and Nutrition, Department of Medicine, University of Maryland School of Medicine, Baltimore, MD.

## Abstract

**Introduction:** Previous studies have suggested that the Amish may experience a relatively high prevalence of Parkinson’s disease (PD) and/or parkinsonian-motor signs.

**Methods:** We assessed the frequency of PD-related motor and no-motor symptoms in a large sample from the Amish Community in Lancaster County, Pennsylvania age ≥ 18 y.

**Results:** Of 430 participants ≥ 60 years, five (1.2%) reported a PD diagnosis, a prevalence similar to estimates in the general older adult populations. Of those without a PD diagnosis, 10.5% reported ≥ 1 and 1.2% ≥ 4 motor symptoms for the nine-item PD screening questionnaire. We also used questionnaires to assess non-motor symptoms. Constipation was reported in 0.7%, and daytime sleepiness in 8.1% of the participants. These frequencies are similar to, or lower than, corresponding frequencies reported in the US National Health and Nutrition Examination Surveys.

**Discussion:** These data neither support a markedly higher PD prevalence in the Lancaster Amish, nor do they show that non-motor symptoms occur with prevalence different that the general US population. It is possible that the Lancaster Amish differ from other US Amish populations in genetics or environmental exposures, or that there were methodological differences between this study and prior ones.

## Introduction

Parkinson’s disease (PD) is marked clinically by motor disability and pathologically by nigrostriatal dopaminergic neuronal loss, but also by extensive non-motor involvement and extensive extra-nigral pathology [1]. The exact etiology for the disease is unknown and probably related to the complex interaction among genetic, environment and lifestyle factors. Several hundred Amish immigrated to the US in the 1700s [2]. The communities are characterized by large family sizes and social cohesiveness. Since Amish marry within their religion, they represent a genetic isolate [3]. They generally follow a simple, agriculture-based lifestyle, and experience low rates of smoking, factors known to be associated with decreased risk for PD.

In a community-based survey of 4,369 Amish in Missouri, Racette et al.[4] reported that 7% of 213 participants older than 60 years met the criteria of clinical diagnosis of PD and 35% had subclinical parkinsonian signs based on Unified PD Rating Scale-Part III motor (UPDRS) exams. Similarly, Cummings et al. [5] reported a prevalence of ∼5% (33/647) for clinical PD in Amish subjects ≥60 years old from Indiana and Ohio. These data suggest older Amish have an unusually high prevalence of PD than the ∼1% generally reported for older adult populations [6]. This would make the Amish an unique population to study novel risk factors for PD.

Because the Amish communities founded in Pennsylvania, Ohio/Indiana, Illinois, and Missouri represent distinct population isolates, we tested if the Lancaster County, PA Amish population also had this markedly higher prevalence of PD symptoms. Because non-motor symptoms, such as constipation and sleep disorders, are now known to be integral part of PD [7], and often occur prior to the onset of typical motor symptom [8, 9], we also ascertained the prevalence of these non-motor symptoms in this investigation.

## Materials and Methods

### Amish Wellness Study

Lancaster County, PA is home to ∼35,000 Old Order Amish (OOA) individuals, of whom approximately one-half are over the age of 21. In 2010, we initiated the Amish Wellness Study to ascertain the entire adult OOA population of Lancaster County for health assessment and bio-banking for future studies. The study was performed at the Amish Research Clinic in Lancaster, PA or in an RV repurposed for performing field-based clinical research in the community. All Amish in Lancaster County aged 18 years and older were eligible for enrollment. Recruitment was carried out by geographic area, with nurses and liaisons going house-to-house with invitations to participate. Individuals agreeing to participate signed a consent were scheduled for a study visit. In the visit, a nurse well-known to the Amish administered the Wellness evaluation. This report is based on up to 3,789 participants who were administered a medical history questionnaire by the research nurse between January 2010 and February 2016 [10]. Although some of the PD-related questions were included at the inception of the Wellness Study, others, as indicted below, were added in subsequent years. The Amish Wellness Study was approved by the University of Maryland Institutional Review Board, and all procedures were done in accord with the Helsinki Declaration of 1975.

### Assessment Tools

#### Assessment of Parkinson’s-related motor symptoms

In 2012 we extended the medical history questionnaire used in the Amish Wellness Study to ascertain prior diagnosis of PD with the question: “Have you ever been diagnosed by a physician as having PD?” If the answer was “yes”, it was recorded as a prevalent PD case. If the answer was “no”, we then continued by administering a nine-item questionnaire (see Supplemental Table1) that was designed by the PD Epidemiology Research Group [11] to provide the first stage of a door-to-door survey for PD.

#### Assessment of Parkinson’s-related non-motor symptoms

We queried study participants by questionnaire for the presence of two non-motor symptoms that have been commonly linked to PD: bowel movement frequency and daytime sleepiness.

##### Bowel movement frequency

Study participants were asked: “How many times a week do you usually have a bowel movement?” We considered ≤ 3 bowel movements per week to be ‘infrequent’, a definition close to that used in the Health Professionals and Nurses’ Health studies [12], the same definition was also used to study constipation as a PD prodromal symptom in the general population in one of our previous reports [13].

##### Daytime Sleepiness

Study participants were asked: “How often did you feel excessively or overly sleepy during the day?” Allowable responses were: “never”, “rarely (<1/week)”, “sometimes (1-2/week)”, “frequent” (3-4/week)”, and “always (5-7/week)”. We defined daytime sleepiness as “always” sleepy during the day, similar to the definition of daytime sleepiness used in the Honolulu Asia Aging Study (“Are you sleepy most of the day”), which shows a significant correlation with PD risk (OR=3.3) [14]. In addition, a similar cut-off was used to study daytime sleepiness in the general population in our previous report [13].

### Statistical Analysis

We computed the prevalence (and 95% confidence intervals) of all symptoms by age group and gender. For motor symptoms, we calculated the prevalence of reporting one, two, and three or more symptoms. For non-motor symptoms, we compared prevalence rates with those previously reported by our group in the US National Health and Nutrition Examination Surveys (NHANES) [13].

## Results

This report is based on questionnaire responses obtained from 3,789 Amish individuals, of whom 1,076 (28%) were ≥ 60 years old and 2,132 (56%) were women. Because the Amish Wellness Study questionnaires evolved over time, the constipation questionnaire was administered to all 3,789 subjects, the PD symptom questionnaire to 2,025 subjects, and the daytime sleepiness questionnaire to only 1,710 subjects.

Among 2,025 subjects who answered the PD questionnaire, 430 were older than 60 years. Five subjects (1.2%) reported having a PD diagnosis, all of whom were ≥70 years old. Four of these were women. The overall frequency of motor symptoms was very low for subjects younger than 60 (< 1%), dramatically increasing after age 60 with 10.5% reporting “yes” ≥ 1, 5.5% ≥ 2, 3.2% ≥ 3, and 1.2% ≥ 4 for the 9-item PD screening questions, respectively (Table 1 and Supplemental Figure 1). Poor balance was the most commonly reported symptom (4.9%), followed by trouble rising from a chair (3.7%), and shakes and trouble with buttons (2.6%). The prevalence of motor symptoms was similar between males and females, although women were more likely to report poor balance (6.5%) and trouble rising from a chair (4.3%).

**Table 1:**
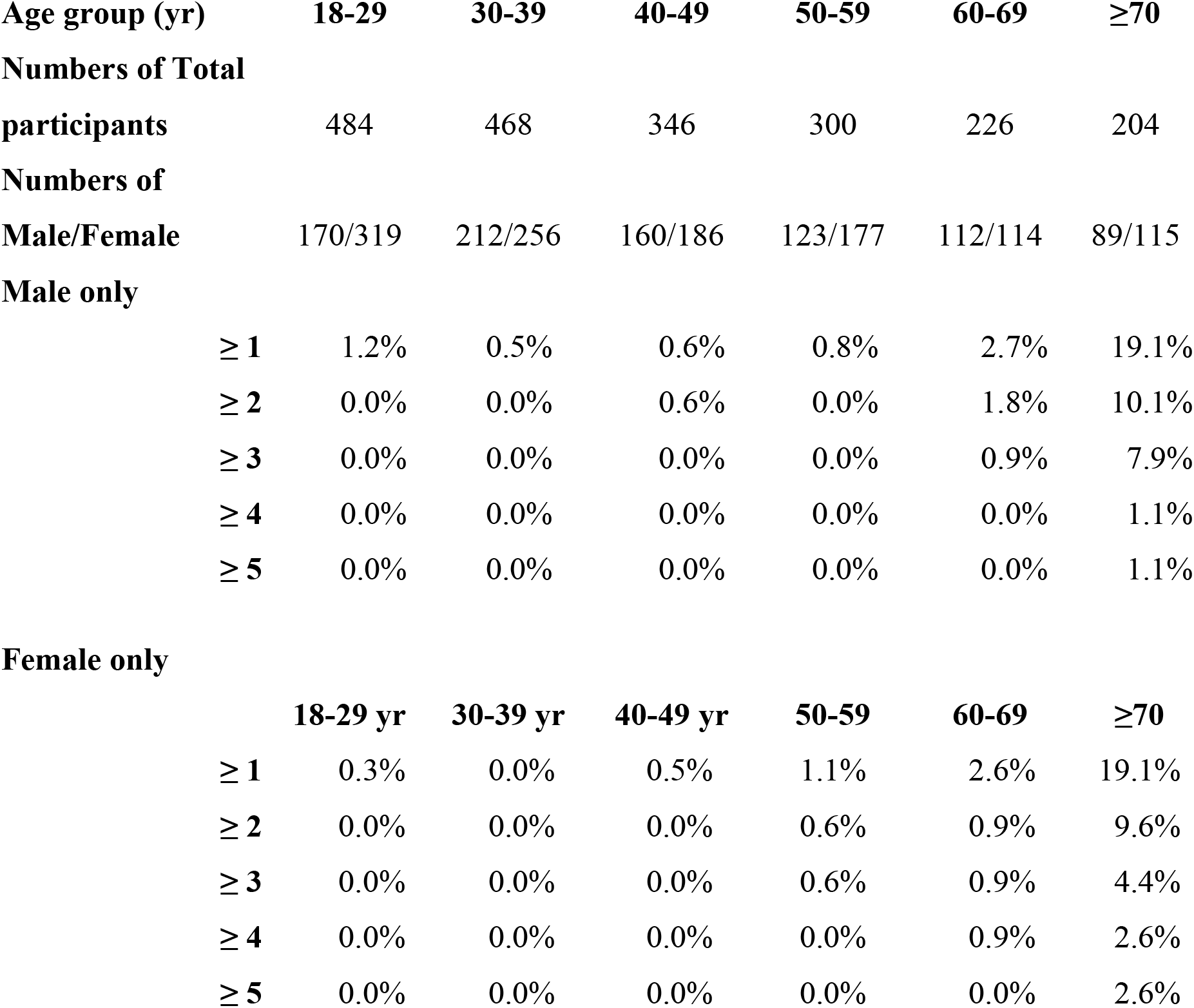
The proportion of Amish men and women reporting ≥ 1, 2, 3, 4, or 5 motor-symptoms according to age.

**Figure.**
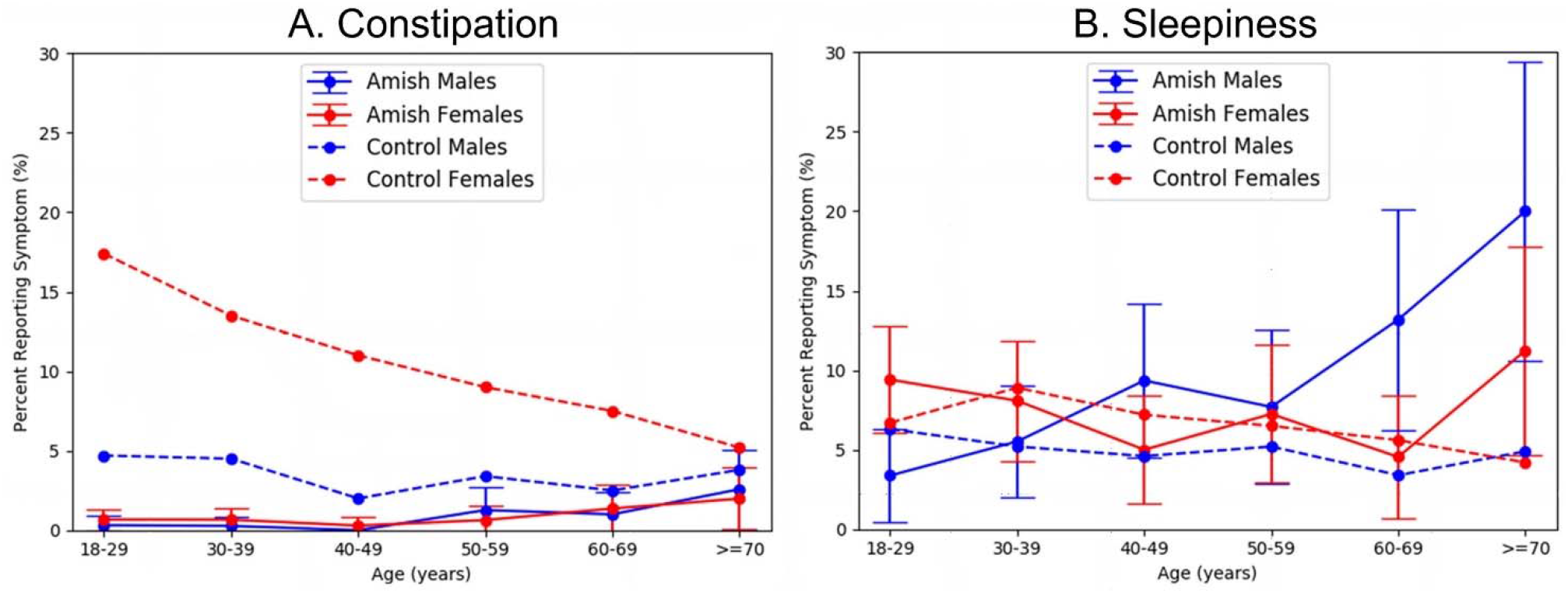
Prevalence of the non-motor symptoms of constipation (A: left) and sleepiness (B: right) reported in the Amish and the National Health and Nutrition Examination Survey, examination cycle 2005-2008 by age group (NHANES data as reported in Chen et al [13]). Error bars represent SD of the mean.

The overall prevalence of constipation was 0.75% (95% CI: 0.47-1.01%), increasing only slightly after age 60 in both men and women. The frequency of daytime sleepiness was high in this population, with an overall rate of 8.1% (95% CI: 6.78%-9.36%).

## Discussion

Contrary to our initial hypothesis based on results from other Amish communities [4, 5], we found that the prevalence of clinical PD diagnoses (1.2%) to be similar to that reported in the general U.S. older adult populations [6]. For those without a PD diagnoses, the prevalence of motor symptoms defined as positive for at least one nine-item motor questionnaire to screen for PD (10.5%) is not higher than the general population based on based on face-face exam in older populations (16%-48%) [15, 16]. Thus, our data do not support a relatively high prevalence of PD in the Lancaster Amish.

This is first study to ascertain the age- and gender-specific prevalence of non-motor symptoms in an Amish community. Specifically when comparing to the United States’ general population as a control (from the NHANES study that we analyzed previously) [13], we did not find higher prevalence of non-motor symptoms in this community. Together, the current data do not support the hypothesis that the Lancaster Amish have a higher prevalence of PD and associated symptoms than the general population. The self-reported infrequent bowel movement in Lancaster Amish was 0.7% as compared to 3.7% reported by NHANES [13] participants. The reason is unclear, but several factors may be involved. The Amish have a generally active lifestyle (e.g. farming, manual labor) that may decrease constipation [10]. Secondly, it is possible that this reflects the population’s dietary habits or microbiome composition. [10] Third, there may be cultural sensitivity to reporting these symptoms [10]. Fourth, it is also possible that this reflects a specific trait in this relatively genetically isolated population. The self-reported sleepiness in our older (>70 yr) Amish population seemed to be higher from that in the general population. It is possible that this finding related to the Amish-specific life styles or behaviors of Amish, but data are too preliminary to make inferences.

Currently, the diagnosis of PD relies on the presence of cardinal motor signs (e.g. tremor at rest, bradykinesia and rigidity) that often are not evident until >50% of nigral dopaminergic neurons have been lost [17, 18]. These, coupled with issues with health care access and bias, have motivated efforts to identify subtle parkinsonian symptoms without clinical diagnoses of PD in the different communities. These efforts have yielded much higher prevalence of Parkinsonian signs. For example, prevalence of subtle parkinsonian signs were reported up to 50% in a cross-sectional study [19], with annual incidence of 3.6-16 incidence in a longitudinal study of older adults [15]. Our study suggests that we have to interpret the recent work with caution because parkinsonian signs do not equal PD, and future studies that can integrate non-motor ascertainment may help to stratify those with parkinsonian signs.

The major strengths of our report include the relatively large sample size, unique Amish population, and a relatively high participation rate of 70%. We do not have data recording why an individual chose not to participate, but the relatively high rate of participation might mitigate concerns about selection bias. The major limitation is the lack of validation of self-reported PD diagnosis and related motor and non-motor symptoms in this population. The responses may have been understated considering the cultural attributes of the Amish (stoicism, gratitude, and uncomplaining). It is also possible that Amish individuals with PD were less likely to participate in our survey.

In summary, this first study to report the prevalence of self-reported PD diagnosis, and PD related motor in the Lancaster County Amish Community does not show the expected higher PD prevalence predicted from earlier studies of other Amish populations. It would be interesting to determine if this is a result either of genetic or lifestyle disparities between the communities, or a consequence of differences in research methodology.

## Data Availability

Data will be made available upon request.

## Author Contributions

The following are the contributions of each author using the following criteria:

1. Research project: A: Conception, B: Organization, C: Execution
2. Data and Statistical analysis: A: Design, B: Execution, C: Review and critique
3. Manuscript preparation: A: Writing the first draft, B: Review and critique

Michael D.F. Goldenberg: 1B, 1C, 2B, 2C, 3A, 3B

Xuemei Huang: 1A, 1B, 1C, 2A, 2C, 3A, 3B.

Honglei Chen: 1A, 1B, 1C, 2A, 2B, 2C, 3B

Lan Kong: 1C, 2A, 2B, 2C, 3B

Teodor T. Postolache: 1A, 2A, 2B, 2C, 3B

John W. Stiller: 1A, 2A, 2B, 2C, 3B Katherine A. Ryan: 1C, 2B, 3B

Mary Pavlovich: 2B, 2C, 3B

Toni I. Pollin: 1A, 1B, 1C, 2A, 2B, 2C, 3B

Alan R. Shuldiner: 1A, 1B, 1C, 2A, 2B, 2C, 3B

Richard B. Mailman: 1A, 3B

Braxton D. Mitchell: 1A, 1B, 1C, 2A, 2B, 2C, 3A, 3B

